# Wastewater sequencing from a rural community enables identification of widespread adaptive mutations in a SARS-CoV-2 Alpha variant

**DOI:** 10.1101/2024.11.11.24316360

**Authors:** Michael J. Conway, Michael P. Novay, Carson M. Pusch, Avery S. Ward, Jackson D. Abel, Maggie R. Williams, Rebecca L. Uzarski, Elizabeth W. Alm

## Abstract

**Background:** Central Michigan University (CMU) participated in a state-wide wastewater monitoring program starting in 2021. One rural site consistently produced higher concentrations of SARS-CoV-2 genome copies. Samples from this site were sequenced retrospectively and exclusively contained a derivative of Alpha variant lineage B.1.1.7 that shed from the same site for 20-28 months.

**Results:** Complete reconstruction of each SARS-CoV-2 open reading frame (ORF) and alignment to an early B.1.1.7 clinical isolate identified novel mutations that were selected in non-structural (nsp1, nsp2, nsp3, nsp4, nsp5/3CLpro, nsp6, RdRp, nsp15, nsp16, ORF3a, ORF6, ORF7a, and ORF7b) and structural genes (Spike, M, and N). These were rare mutations that have not accumulated in clinical samples worldwide. Mutational analysis revealed divergence from the reference Alpha variant lineage sequence over time. We present each of the mutations on available structural models and discuss the potential role of these mutations during a chronic infection.

**Conclusions:** This study further supports that small wastewater treatment plants can enhance resolution of rare events and facilitate reconstruction of viral genomes due to the relative lack of contaminating sequences and identifies mutations that may be associated with chronic infections.

## 1. Introduction

Wastewater monitoring has become a firmly established public health tool since the COVID-19 pandemic. Wastewater monitoring programs have helped identify potential outbreaks within communities and individual buildings, they can track variants of concern, and they are being leveraged for new emerging infectious diseases (1–12). The goal of wastewater monitoring is to provide complementary data to public health agencies so that they can make informed decisions to mitigate infectious disease transmission.

The State of Michigan Department of Health and Human Services (MDHHS) initiated a wastewater monitoring program in 2021. The program included partnerships between academic laboratories and regional public health departments that spanned large and small metropolitan areas and rural areas in both lower and upper peninsulas. Central Michigan University (CMU) formed a partnership with the Central Michigan District Health Department (CMDHD). This partnership provided an opportunity to look at the dynamics of SARS-CoV-2 at a regional public university and in the surrounding small metropolitan and rural communities (13). We identified ten on-campus sewer sites and nine off-campus wastewater treatment plants (WWTPs) to sample on a weekly basis.

Sampling began in July 2021, which was at least seven months after emergence of the Alpha variant (B.1.1.7) in Michigan. The Alpha variant first appeared in North America in late November 2020 and became the predominant SARS-CoV-2 variant by the end of March 2021 (14). It became clear that our smallest WWTP (estimated population served: 851) consistently produced higher concentrations of SARS-CoV-2 genome copies. Samples taken from this site from 2021-2023 were retrospectively sequenced and many contained sequences that corresponded to an Alpha variant lineage B.1.1.7. We reconstructed the Spike gene and identified 37 mutations that accumulated in the RBD and NTD (15). Here, we use the same set of data to provide a complete reconstruction of each SARS-CoV-2 open reading frame (ORF). Alignment of each ORF to an early B.1.1.7 clinical isolate identified novel mutations that were selected in non-structural (nsp1, nsp2, nsp3, nsp4, nsp5/3CLpro, nsp6, RdRp, nsp15, nsp16, ORF3a, ORF6, ORF7a, and ORF7b) and structural genes (Spike, M, and N). Each of these mutations were present in less than 2% of clinical samples present in GenBank and the sequence read archive (SRA) from Dec 2023 to Jun 2024. These were rare mutations, yet three were previously associated with immunodeficiency, adaptation to remdesivir, and reinfection of a hospital worker (16–20). Temporal mutational analysis revealed divergence from the reference Alpha variant lineage sequence over time. Each mutation was mapped onto available structural models, and we discuss the potential significance of these changes during a chronic SARS-CoV-2 infection.

This manuscript provides further support that wastewater monitoring in small metropolitan and rural communities is an opportunity to identify novel variants and reconstruct whole genomes due to lower contamination with unrelated sequences. It is important to note that the reconstruction strategy that was used will incorporate contaminating variant sequences if they are present. These data also support that humans can chronically shed SARS-CoV-2 for close to two years. Considering the low prevalence of these mutations in clinical samples, chronic shedding of SARS-CoV-2 is likely a rare event that leads to accumulation of adaptive mutations. Identifying mutations associated with chronic infection may be useful to diagnose individuals who have persistent disease and to assist in the selection of appropriate treatment.

## 2. Materials and methods

### 2.1. Selection of sample sites

Central Michigan University (CMU) is a public research university in the City of Mt. Pleasant, Isabella County, Michigan, with an average population during the 2021-2022 academic year of 13,684 students and staff. Ten sample sites were selected on campus that collected wastewater downstream from most campus buildings, including residential halls, apartments, and academic/administrative buildings. The waste stream at these sites includes a mixture of wastewater from CMU and upstream residential areas in the City of Mt. Pleasant. Nine off-campus sites throughout the jurisdictions of the Central Michigan District Health Department (CMDHD) and Mid-Michigan District Health Department (MMDHD) were selected (13), which included the City of Mt. Pleasant, Union Township, City of Alma, City of Clare, City of Evart, three Houghton Lake townships, and Village of Marion wastewater treatment plants (WWTPs). These locations represent various land uses and population densities including urban, rural, and suburban areas, providing a large footprint of SARS CoV-2 virus shedding in Central Michigan.

### 2.2. Wastewater collection

Since July 2021, wastewater samples (500–1000 mL) were collected once each week on either Monday or Tuesday from ten sanitary sewer sites and nine WWTP influent streams (after grit removal). Sanitary sewer grab samples consisted of wastewater flowing from university dormitories and buildings and the surrounding community. Influent to WWTPs were collected as grab samples or 24-hour composite samples. Samples were held at 4°C no more than 48 hours before analysis (13).

### 2.3. Virus concentration and RNA extraction

The protocol described by Flood et al. 2021 and adopted by the Michigan wastewater monitoring network was used to concentrate virus from samples and extract viral RNA (13, 53). Briefly, 100 mL wastewater or water as a negative control was mixed with 8% (w/v) molecular biology grade PEG 8000 (Promega Corporation, Madison WI) and 0.2 M NaCl (w/v). The sample was mixed slowly on a magnetic stirrer at 4 °C for 2-16 hours. Following overnight incubation, samples were centrifuged at 4,700×g for 45 min at 4 °C. The supernatant was then removed, and the pellet was resuspended in the remaining liquid, which ranged from 1-3 mL. All sample concentrates were aliquoted and stored at −80 °C until further processing. Viral RNA was extracted from concentrated wastewater using the Qiagen QIAmp Viral RNA Minikit according to the manufacturer’s protocol with previously published modifications (Qiagen, Germany) (53). In this study, a total of 200 µl of concentrate was used for RNA extraction resulting in a final elution volume of 80 µl. Extracted RNA was stored at −80 °C until analysis. A wastewater negative extraction control was included. To derive recovery efficiencies for each sample site, samples were inoculated with 10^6^ gene copies (GC)/mL Phi6 bacteriophage (Phi6) prior to the addition of PEG and NaCl. Wastewater samples were mixed, and a 1 mL sample was reserved and stored at −80 °C. RNA was extracted as stated above.

### 2.4. Detection and quantification of SARS-CoV-2

A one-step RT-ddPCR approach was used to determine the copy number/20 µL of SARS-CoV-2, and data were converted to copy number/100 mL wastewater for N1 and N2 targets using the method published by Flood et al., 2001 (53). All the primers and probes used in this study were published previously (13). Droplet digital PCR was performed using Bio-Rad’s 1-Step RT-ddPCR Advanced kit with a QX200 ddPCR system (Bio-Rad, CA, USA). Each reaction contained a final concentration of 1 × Supermix (Bio-Rad, CA, USA), 20 U μL^-1^ reverse transcriptase (RT) (Bio-Rad, CA, USA), 15 mM DTT, 900 nmol l^-1^ of each primer, 250 nmol l^-1^ of each probe, 1 µL of molecular grade RNAse-free water, and 5.5 μL of template RNA for a final reaction volume of 22 μL (13, 53–55). RT was omitted for DNA targets. Droplet generation was performed by microfluidic mixing of 20 μL of each reaction mixture with 70 μL of droplet generation oil in a droplet generator (Bio-Rad, CA, USA) resulting in a final volume of 40 μL of reaction mixture-oil emulsions containing up to 20,000 droplets with a minimum droplet count of > 9000. The resulting droplets were then transferred to a 96-well PCR plate that was heat-sealed with foil and placed into a C1000 96-deep-well thermocycler (Bio-Rad, CA, USA) for PCR amplification using the following parameters: 25 °C for 3 min, 50 °C for 1 h, 95 °C for 10 min, followed by 40 cycles of 95 °C for 30 s and 60 °C for 1 min with ramp rate of 2 °C/s 1 followed by a final cycle of 98 °C for 10 min. Following PCR thermocycling, each 96-well plate was transferred to a QX200 Droplet Reader (Bio-Rad, CA, USA) for the concentration determination through the detection of positive droplets containing each gene target by spectrophotometric detection of the fluorescent probe signal. All analyses were run in triplicate for each marker. To derive recovery efficiencies for each sample site, Phi6-spiked pre- and post-PEG concentration RNA samples were used to quantify Phi6 copy number using the previously published primers and probes (13). The degree of PCR inhibition was also quantified in each sample by spiking 10 μL of 10^5^ GC/ml Phi6 in a sample’s Buffer AVL, including positive controls that lacked wastewater.

### 2.5. Data analysis

All SARS-CoV-2 gene data were converted from GC per 20 µL reaction to GC per 100 mL wastewater sample before analysis (13, 53). Non-detects (ND) were assigned their individual sample’s limit of detection for the purposes of data reporting, although any weekly on-campus or off-campus samples whose values matched the theoretical limit of detection were removed prior to statistical analysis. The limit of detection was calculated for each individual sample based on both the molecular assays’ theoretical detection limits (i.e., 3 positive droplets for RT-ddPCR; the lowest standard curve concentration for RT-qPCR) and the concentration factor of each processing method examined. All wastewater data were reported to MDHHS and uploaded to the Michigan COVID-19 Sentinel Wastewater Epidemiological Evaluation Project (SWEEP) dashboard (https://www.michigan.gov/coronavirus/stats/wastewater-surveillance/dashboard/sentinel-wastewater-epidemiology-evaluation-project-sweep).

### 2.6. Sequencing

RNA was shipped to GT Molecular (Fort Collins, CO) on dry ice. Library preparation was done using GT Molecular’s proprietary method, which utilized ARTIC 4.1 primers for SARS-CoV-2 amplicon generation (https://artic.network/ncov-2019). Amplicons were pooled and sequenced on a Miseq using 2x150bp reads. FASTQ files were analyzed using GT Molecular’s bioinformatics pipeline, and variant-calling was performed using a modified and proprietary version of Freyja (56). FASTQ files for each sample listed are available in the NCBI SRA database (Submission ID: SUB13897431; BioProject ID: PRJNA1027333) (15).

### 2.7. Complete reconstruction and identification of novel mutations

FASTQ files from 10-26-21 (SAMN37791375), 11-9-21 (SAMN37791376), 9-12-22 (SAMN37791379), 3-13-23 (SAMN37791380), 4-24-23 (SAMN37791382), and 5-1-23 (SAMN37791383) contained reads that spanned each SARS-CoV-2 open reading frame (ORF), they lacked contamination with other variants of concern based on variant calling, and they had high relative abundance of the Alpha variant lineage B.1.1.7 derivative (Table 1) (15). This allowed for reconstruction of consensus genes for each of the above wastewater samples. Specifically, we uploaded FASTA-formatted .txt files into Galaxy (https://usegalaxy.org/) that represented each reference gene. Reference genes were constructed from an early consensus Alpha variant lineage B.1.1.7 Michigan clinical isolate submitted on 1-26-21 (GenBank: MW525061.1; Accession: MW525061). We then uploaded each of the paired-end FASTQ files for each wastewater sample. The Bowtie2 program was used to map reads against each reference sequence, creating individual .bam files per sample. The default setting was used for analysis. The Convert Bam program was then used to convert .bam files to FASTA multiple sequence alignments. Multiple sequence alignment files were uploaded to MEGA (https://www.megasoftware.net/) and converted to amino acid sequence. The consensus amino acid sequence from each of these samples was manually reconstructed and then aligned with the reference gene. Mutations that were present in wastewater samples but not the reference clinical sample were characterized as novel mutations. The total number of reads that aligned to each reference gene were determined in MEGA and FastQC was used to quantify read length and the number of poor-quality sequences (Supplementary Table 1). At least 3 reads were present for each amino acid.

**Table 1.** GT Molecular Variant Calling.

| Location code | Sample date | VOC (%)<sup>a</sup> | Lineage(s) (%)<sup>b</sup> |
| --- | --- | --- | --- |
| VM | 10-26-21 | Alpha (94.2) | Q.3 (94.2) |
| VM | 11-9-21 | Alpha (94.9) | Q.4 (94.7) |
| VM | 9-12-22 | Alpha (97.3) | Q.4 (96.9) |
| VM | 3-13-23 | Alpha (98.0) | Q.4 (65.5) |
| VM | 4-24-23 | Alpha (97.9) | Q.4 (89.7) |
| VM | 5-1-23 | Alpha (97.4) | Q.4 (97.1) |

*a, Relative abundance of variants of concern (VOC) as a percentage*
*b, Relative abundance of VOC lineages as a percentage*

### 2.8. Novel mutation hotspot analyses

We identified novel mutations as described above. We then tracked the percent prevalence of novel mutations in wastewater samples that were positive for the Alpha variant lineage. Specifically, we uploaded FASTA-formatted .txt files into Galaxy (https://usegalaxy.org/) that represented the SARS-CoV-2 reference genes. We then uploaded each of the paired-end FASTQ files for each wastewater sample. The Bowtie2 program was used to map reads against the reference sequence. The default setting was used for analysis. The Convert Bam program was then used to convert .bam files to FASTA multiple sequence alignments. Multiple sequence alignment files were uploaded to MEGA (https://www.megasoftware.net/) and converted to amino acid sequence for open-reading frame analysis. Novel mutations were identified manually, and the column of reads were copied and pasted into Excel. The column was selected, and the Analyze Data tool was selected to calculate the percent prevalence of the novel mutations. This was repeated for each novel mutation across all samples positive for Alpha variant lineage and the percent prevalence data was represented in a heatmap. Novel mutations were mapped onto 2-D representations of proteins and the 3-D protein structures when available using UCSF Chimera (57).

## 3. Results

### 3.1 Chronic shedding of an Alpha variant lineage at a rural WWTP

Wastewater samples were collected between July 2021 and June 2023 and SARS-CoV-2 genome copies per 100 mL wastewater were determined each week and reported to MDHHS (13, 15). One site was notable for higher peaks of virus shedding, which culminated in a peak that was 4 logs higher than the mean for all sites, although high peaks of activity were observed since 9-21-21 (15). In order to identify the SARS-CoV-2 variant responsible for this activity, RNA extracted from stored wastewater concentrates was shipped to GT Molecular (Fort Collins, CO) and a next generation sequencing (NGS) and variant calling pipeline was employed. RNA from the site of interest and neighboring sites were analyzed as a control. The site of interest contained high relative abundance of Delta variant lineage AY.25.1 at the first time point tested (i.e., 9-21-21) (15). This corresponded to the beginning of the Delta variant wave in Central Michigan (13). The site of interest began shedding the Alpha variant lineage during the next two time points tested (10-26-21 and 11-9-21) (15). This was preceded by sequencing data from clinical samples, which revealed 16 Alpha variant lineage Q.3 isolates collected from 2-18-21 to 7-9-21 (15). The site of interest had high relative abundance of Omicron variant lineages during the next two time points tested (i.e., 3-14-22 and 4-25-22) (15). This corresponded to the end of the first Omicron wave in Central Michigan (13). The Alpha variant lineage became the dominant isolate in all remaining wastewater samples from the site of interest in all 2022 and 2023 samples tested, with relative abundance ranging from 47.1-98.0% (15). Specifically, for the samples analyzed in the current manuscript (i.e., 10-26-2021, 11-9-2021, 9-12-2022, 3-13-23, 4-24-23, and 5-1-23), the alpha variant represented 94.2-98% of amplicons. Based on this information, it was possible to reconstruct alpha variant genomes without significant contamination with other variants (Supplementary Figure 1). Other sites contained Omicron variant lineages BG.5, XBB.1.5, XBB.1.5.23, XBB.1.28, XBB.1.5.1, XBB.1.5.17, XBB.1.5.49, and Delta variant lineage DT.2 at varying relative abundance during the same sampling period (15).

### 3.2. Accumulation of novel mutations

We reasoned that chronic shedding of SARS-CoV-2 would lead to accumulation of novel mutations that do not align with sequences identified in most clinical and wastewater samples, which mostly result from acute infection. This hypothesis was supported by our previous analysis of Spike, which identified 37 novel mutations that accumulated during this time frame (15). Alignment of reconstructed consensus genes with a reference Alpha variant lineage clinical sequence revealed that non-Spike proteins had 35 novel mutations in the 5-1-2023 sample including mutations in nsp1 (1), nsp2 (1), nsp3 (8), nsp4 (4), nsp5/3CLpro (2), nsp6 (2), RdRp (2), nsp15 (2), nsp16 (1), ORF3a (4), M (2), ORF6 (1), ORF7a (1), ORF7b (1), and N (3) (Fig. 1). Each mutation was analyzed using the NCBI Virus SARS-CoV-2 Variant Overview “Search GenBank + SRA Data by Mutation” tool. This analysis identified the total records for each mutation in GenBank and SRA databases during the time frame of Dec 2023 to Jun 2024. Importantly, this time frame was 7 to 13 months after the last wastewater sample was acquired. Very few records were identified for each of these mutations and some mutations lacked records altogether (Table 2). We expanded the mutational analysis by quantifying the percent prevalence of each of the 35 novel mutations identified in the 5-1-2023 sample across all wastewater samples that were positive for the Alpha variant lineage. A heatmap showed that these mutations accumulated and became dominant within the population over time, while also retaining diversity at each position (Fig. 2).

**Figure 1.**
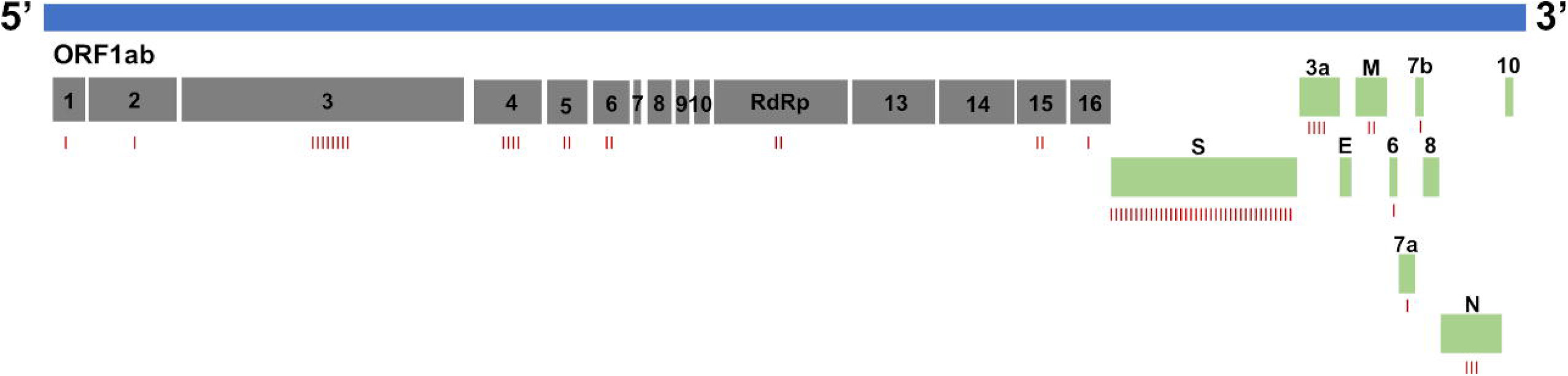
SARS-CoV-2 genome map with novel mutations identified as red dashes under each open reading frame (ORF): nsp1 (1), nsp2 (1), nsp3 (8), nsp4 (4), nsp5/3CLpro (2), nsp6 (2), RdRp (2), nsp15 (2), nsp16 (1), S (37), ORF3a (4), M (2), ORF6 (1), ORF7a (1), ORF7b (1), and N (3).

**Figure 2.**
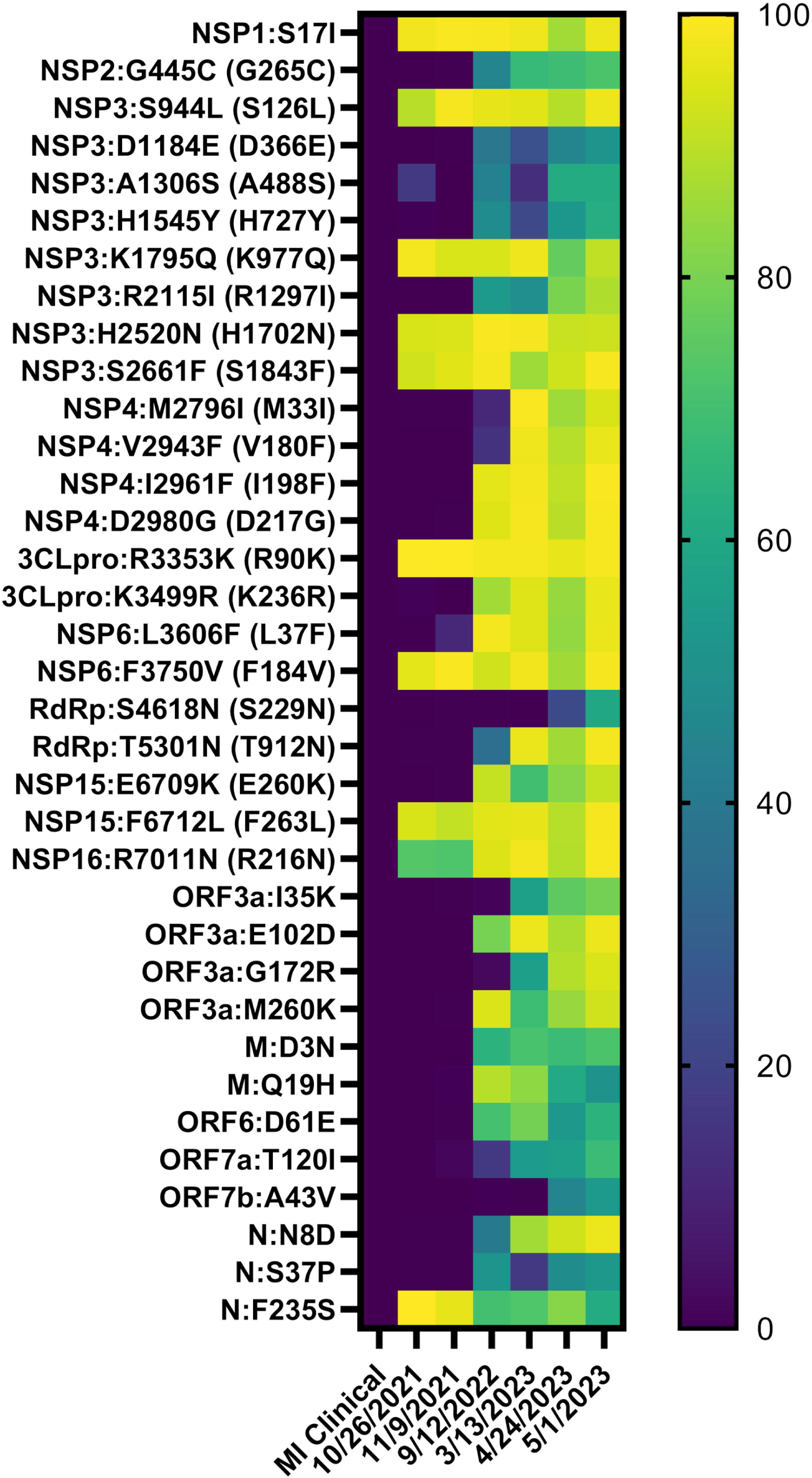
Heatmap showing the percent prevalence of novel non-Spike mutations in each wastewater sample that was positive for the Alpha variant lineage. ORF1ab mutations outside of parentheses were determined from the first amino acid in nsp1. ORF1ab mutations inside of parentheses were determined from the first amino acid of each protein.

**Table 2.** Total Records of Non-Spike Mutations Identified in a Chronic SARS-CoV-2 Alpha Variant.

| MUTATION | SRA | GenBank | Unique Samples |
| --- | --- | --- | --- |
| NSP1:S17I | 1 | 0 | 1 |
| NSP2:G445C (G265C)<sup>a</sup> | 6 | 20 | 21 |
| NSP3:S944L (S126L) | 60 | 148 | 111 |
| NSP3:D1184E (D366E) | 6 | 8 | 6 |
| NSP3:A1306S (A488S) | 0 | 0 | 0 |
| NSP3:H1545Y (H727Y) | 1 | 4 | 2 |
| NSP3:K1795Q (K977Q) | 2 | 11 | 8 |
| NSP3:R2115I (R1297I) | 13 | 1 | 13 |
| NSP3:H2520N (H1702N) | 0 | 0 | 0 |
| NSP3:S2661F (S1843F) | 19 | 23 | 25 |
| NSP4:M2796I (M33I) | 33 | 39 | 41 |
| NSP4:V2943F (V180F) | 1 | 2 | 1 |
| NSP4:I2961F (I198F) | 0 | 0 | 0 |
| NSP4:D2980G (D217G) | 11 | 10 | 11 |
| 3CLpro:R3353K (R90K) | 0 | 0 | 0 |
| 3CLpro:K3499R (K236R) | 1 | 4 | 4 |
| NSP6:L3606F (L37F) | 714 | 984 | 977 |
| NSP6:F3750V (F184V) | 2 | 6 | 6 |
| RdRp:S4618N (S229N) | 13 | 29 | 18 |
| RdRp:T5301N (T912N) | 0 | 0 | 0 |
| NSP15:E6709K (E260K) | 2 | 2 | 2 |
| NSP15:F6712L (F263L) | 6 | 8 | 8 |
| NSP16:R7011N (R216N) | 0 | 0 | 0 |
| ORF3a:I35K | 42 | 56 | 46 |
| ORF3a:E102D | 1 | 2 | 2 |
| ORF3a:G172R | 0 | 1 | 0 |
| ORF3a:M260K | 11 | 17 | 15 |
| M:D3N | 13 | 14 | 19 |
| M:Q19H | 0 | 0 | 0 |
| ORF6:D61E | 0 | 0 | 0 |
| ORF7a:T120I | 34 | 72 | 63 |
| ORF7b:A43V | 43 | 68 | 65 |
| N:N8D | 2 | 3 | 2 |
| N:S37P | 28 | 48 | 48 |
| N:F235S | 0 | 0 | 0 |

<sup>a</sup> ORF1ab mutations outside of parentheses were determined from the first amino acid in *nsp1*. ORF1ab mutations inside of parentheses were determined from the first amino acid of each protein and were required for this analysis.

### 3.3. Nsp1

One novel mutation accumulated in nsp1: S17I. S17I resides in the N-terminal domain (NTD) (Figure 3A). It was found in 1 online record (Table 2). The impact of this mutation on protein function and immune evasion is unknown.

**Figure 3.**
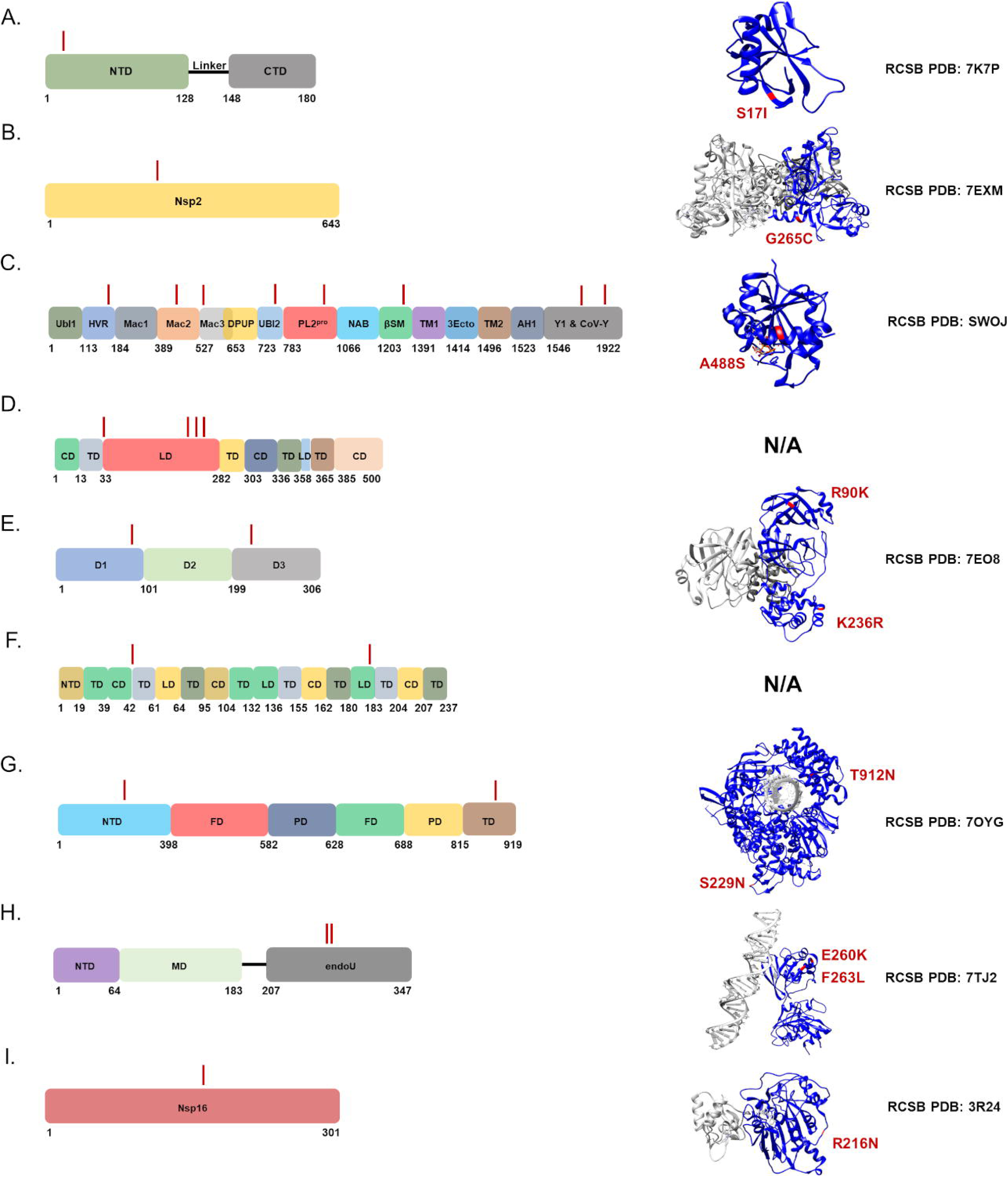
Novel mutations present in ORF1ab genes were mapped onto 2-D and 3-D protein models: (A) nsp1, (B) nsp2, (C) nsp3, (D) nsp4, (E) nsp5/3CLpro, (F) nsp6, (G) RdRp, (H) nsp15, and (I) nsp16. Structures were rendered using UCSF Chimera and RCSB PDB numbers were provided (58). Individual protein domains were indicated in blue and multimers/other molecules were indicated as gray. Mutations were highlighted and noted in red.

### 3.4. Nsp2

One novel mutation accumulated in nsp2: G445C (G265C). ORF1ab mutations outside of parentheses were determined from the first amino acid in nsp1. ORF1ab mutations inside of parentheses were determined from the first amino acid of each protein. Both variations were searched in the literature. G445C (G265C) was reported in India and found in a total of 26 online records (Table 2) (21). The impact of this mutation on protein function and immune evasion is unknown.

### 3.5. Nsp3

Eight novel mutations accumulated in nsp3: S944L (S126L), D1184E (D366E), A1306S (A488S), H1545Y (H727Y), K1795Q (K977Q), R2115I (R1297I), H2520N (H1702N), and S2661F (S1843F). S944L (S126L) resides in the hypervariable region (HVR), D1184E (D366E) resides in macrodomain II (Mac2), A1306S (A488S) resides in macrodomain III (Mac3), H1545Y (H727Y) resides in the ubiquitin-like domain 2 (Ubl2), K1795Q (K977Q) resides in the PL2^pro^ domain, R2115I (R1297I) resides within the betacoronavirus-specific marker (βSM), and H2520N (H1702N) and S2661F (S1843F) reside in the Y1 and CoV-Y domains (Figure 3C). S944L (S126L), A1306S (A488S), and S2661F (S1843F) were reported in Asian isolates and found in a total of 208, 0, and 42 online records, respectively (Table 2) (22–25). H1545Y (H727Y) was reported in Italy and found in a total of 5 online records (Table 2) (26). K1795Q (K977Q) was reported in Brazil, was present at a higher frequency in immunodeficient patients, and found in a total of 13 online records (Table 2) (27, 28). R2115I (R1297I) was reported in Moldova and found in a total of 14 online records (29). D1184E (D366E) and H2520N (H1702N) have not been previously reported and were found in a total of 14 and 0 online records, respectively. The impact of these mutations on nsp3 protein function and immune evasion is unknown.

### 3.6. Nsp4

Four novel mutations accumulated in nsp4: M2796I (M33I), V2943F (V180F), I2961F (I198F), and D2980G (D217G). M2796I (M33I) resides in a transmembrane domain (TD), and V2943F (V180F), I2961F (I198F), and D2980G (D217G) reside in the luminal domain (LD) (Figure 3D). M2796I (M33I) was reported in the Middle East and computational analysis suggested that the mutation causes secondary structure changes converting an alpha helix to a beta sheet, which may impact interaction between nsp3 and nsp4 (30, 31). D2980G (D217G) and M2796I (M33I) were reported in Asian isolates and were found in a total of 21 and 72 online records, respectively (32, 33). V2943F (V180F) and I2961F (I198F) have not been previously reported and were found in a total of 3 and 0 online records, respectively. The impact of these mutations on protein function and immune evasion are unknown.

### 3.7. Nsp5/3CLpro

Two novel mutations accumulated in nsp5: R3353K (R90K) and K3499R (K236R). R3353K (R90K) resides in domain 1 (D1) and K3499R (K236R) resides in D3 (Figure 3E). R3353K (R90K) was not previously reported and was found in 0 online records (Table 2). K3499R (K236R) was reported in Indian isolates and was found in a total of 5 online records (Table 2) (32). The impact of these mutations on protein function and immune evasion are unknown.

### 3.8. Nsp6

Two novel mutations accumulated in nsp6: L3606F (L37F) and F3750V (F184V). L3606F (L37F) resides in a transmembrane domain (TD) and F3750V (F184V) resides in a luminal domain (LD) (Figure 3F). L3606F (L37F) was reported in Asian isolates and was found in a total of 1,698 (2%) online records (Table 2) (34). Previous research found that L3606F (L37F) reduced nsp6’s interaction with ATP6AP1. This allowed for lysosomal acidification to proceed normally, which prevented activation of the NLRP3 inflammasome pathway. The investigators noted that this mutation reduced SARS-CoV-2 fitness, and that this may be why the mutation is not present in circulating variants of concern (35, 36). F3750V (F184V) was reported in a basic science study as an adaptive mutation that arose in ferrets and *in vitro* during treatment with remdesivir (16, 17). This mutation was not reported in clinical sequences but was present in a total of 8 online records (Table 2).

### 3.9. RdRp

Two novel mutations accumulated in RdRp: S4618N (S229N) and T5301N (T912N). S4618N (S229N) resides in the NTD and T5301N (T912N) resides in the thumb domain (TD) (Figure 3G). S4618N (S229N) was not previously reported and was present in a total of 42 online records (Table 2). T5301N (T912N) was reported in Italy and was present in a total of 0 online records (Table 2) (37). The impact of these mutations on protein function and immune evasion are unknown.

### 3.10. Nsp15

Two novel mutations accumulated in nsp15: E6709K (E260K) and F6712L (F263L). Both E6709K (E260K) and F6712L (F263L) reside in the nuclease EndoU domain (Figure 3H). E6709K (E260K) was reported in Italy and was present in a total of 4 online records (Table 2). This mutation was associated with an increased frequency of mortality (38). F6712L (F263L) was not previously reported and was present in a total of 14 online records (Table 2). The impact of these mutations on protein function and immune evasion are unknown.

### 3.11. Nsp16

One novel mutation accumulated in nsp16: R7011N (R216N). R7011N (R216N) was reported in Brazil and was present in a total of 0 online records (Table 2). This mutation was associated with reinfection of a healthcare worker (19). The impact of this mutation on protein function and immune evasion is unknown.

### 3.12. ORF3a

Four novel mutations accumulated in ORF3a: I35K, E102D, G172R, and M260K. I35K resides in the N-terminal domain (NTD), E102D resides in the transmembrane domain (TD), and both G172R and M260K reside in the C-terminal domain (CTD) (Figure 4A). I35K and E102D have not been previously reported. G172R and M260K were previously reported in variants of concern (39–42). The impact of these mutations on ORF3a protein function and immune evasion is unknown.

**Figure 4.**
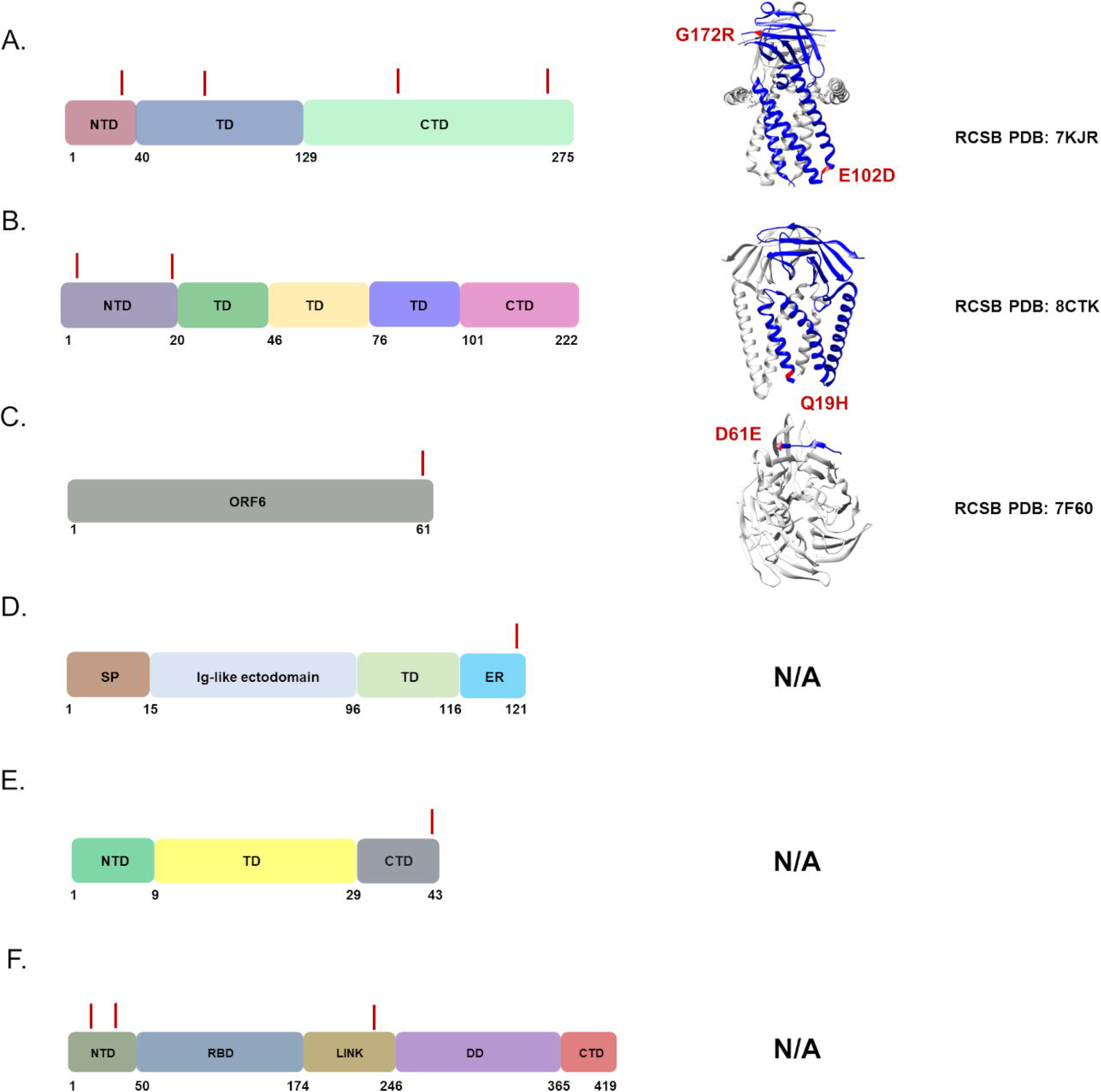
Novel mutations present in ORF genes were mapped onto 2-D and 3-D protein models. (A) ORF3a, (B) M, (C) ORF6, (D) ORF7a, (E) ORF7b, and (F) N. Structures were rendered using UCSF Chimera and RCSB PDB numbers were provided (58). Individual protein domains were indicated in blue and multimers/other molecules were indicated as gray. Mutations were highlighted and noted in red.

### 3.13. M

Two novel mutations accumulated in M: D3N and Q19H. Both mutations reside in the N-terminal domain (NTD), which are surface exposed (Figure 4B). D3N was previously identified in BA.5 strains and may result in N-myristoylation possibly impacting membrane integrity, post-translational modification, and immune evasion (43, 44). Q19H has not been previously reported but based on its location it may have similar functional consequences as D3N.

### 3.14. ORF6

One novel mutation accumulated in ORF6: D61E. This is the last residue in the open reading frame. D61E has not been previously reported. The impact of this mutation on ORF6 protein function and immune evasion is unknown.

### 3.15. ORF7a

One novel mutation accumulated in ORF7a: T120I. This is the second to last C-terminal residue and resides in the endoplasmic reticulum retention sequence (Figure 4D). T120I has been previously identified in variants of concern (45). The impact of this mutation on ORF7a protein function and immune evasion is unknown.

### 3.16. ORF7b

One novel mutation accumulated in ORF7b: A43V. This is the last residue in the C-terminal domain (Figure 4E). A43V has not been previously reported. The impact of this mutation on ORF7b protein function and immune evasion is unknown.

### 3.17. N

Three novel mutations accumulated in the N gene: N8D, S37P, and F235S. N8D resides in the N-terminal domain (NTD) and both S37P and F235S reside in the linker sequence between the known functional domains (Figure 4F). N8D was previously identified in B.1.1.7 strains (46, 47). S37P was previously identified in SARS-CoV-2 isolates (48, 49). F235S has not been previously reported. The impact of these mutations on N protein function and immune evasion is unknown.

## 4. Discussion

Retrospective analysis of wastewater data revealed that one rural site produced consistently higher concentrations of SARS-CoV-2 copy numbers. NGS sequencing revealed that this site began shedding an Alpha variant lineage by October 2021 and that this continued to at least May 2023. Clinical sequence data revealed that Alpha variant lineage Q.3/Q.4 was present in Michigan between February to July 2021. This preceded the start of wastewater surveillance in central Michigan and our first detection of the Alpha variant lineage in wastewater by 3-8 months. It is unclear how many individuals were originally infected with this lineage at the site of interest, and it is unclear how many individuals continued to shed the virus into the sewer shed. However, due to the small population served at this rural WWTP, our October 2021 Alpha variant lineage reconstruction likely represents a chronic infection that lasted for 2-7 months. At this stage of the chronic infection, the Alpha variant lineage already accumulated 9 novel mutations in the Spike gene and 10 novel mutations spread across nsp1, nsp3, nsp5, nsp6, nsp15, nsp16, and N (15).

Eighteen months later, the Alpha variant lineage shed from this site had 72 novel non-structural (nsp1, nsp2, nsp3, nsp4, nsp5/3CLpro, nsp6, RdRp, nsp15, nsp16, ORF3a, ORF6, ORF7a, and ORF7b) and structural (Spike, M, and N) mutations (15). Spike mutations present in these wastewater samples have been described previously. Many of the Spike mutations were associated with experimental evidence suggesting they promoted immune evasion, and three mutations were previously found in immunocompromised patients (15). Similarly, nsp3 K1795Q (K977Q) was present at a higher frequency in immunodeficient patients, nsp6 F3750V (F184V) was an adaptive mutation that accumulated in the presence of remdesivir *in vitro* and *in vivo*, and nsp16 R7011N (R216N) was present in a reinfected healthcare worker (16, 17, 19, 28). Considering that 8% of the novel mutations in Spike and non-Spike genes were associated with known persistent infections, we can assume that many of the identified mutations are selected during chronic infection.

Without experimental evidence, it is impossible to know the precise role of each mutation during chronic infection, although we can infer based on known structural and functional information for each protein, in addition to serological data in the human host. For instance, the two mutations present in M protein (D3N and Q19H) reside in the N-terminal domain (NTD) and are surface exposed (50, 51). Previous research has identified D3N in BA.5 strains and investigators suggested that the mutation may result in N-myristoylation possibly impacting membrane integrity, post-translational modification, and immune evasion (43, 44). Similar to Spike, antibodies directed to M are generated during infection, persist for at least a year after infection, and generate a similar level of reactivity as immunodominant linear epitopes (50–52). It’s possible that during a chronic infection SARS-CoV-2 optimizes Spike and M to evade adaptive immunity.

These data provide evidence that, while rare, an individual can be chronically infected with SARS-CoV-2 over many months and possibly a few years. It is possible that multiple individuals contributed to the persistence of this variant of concern. During this time, SARS-CoV-2 can accumulate many mutations in Spike and non-Spike genes. Some of these mutations have been found in persistent infections. Further research is needed to determine which of these mutations are predictive of chronic infection and if they can be used as a biomarker in individuals with persistent disease and leveraged to tailor selection or development of pharmaceutical therapies. This study also shows that small WWTPs can enhance the resolution of rare biological events and allow for total reconstruction of viral genomes and their corresponding proteins.

## Declarations

### Ethics approval and consent to participate

Not applicable

### Consent for publication*

Not applicable

### Availability of data and materials

FASTQ files for each sample are available in the NCBI SRA database (Submission ID: SUB13897431; BioProject ID: PRJNA1027333).

### Authors’ contributions

MJC wrote the manuscript and directed the wastewater monitoring activities, MPN reconstructed ORFs, CMP analyzed FASTQ data and developed the heatmap, ASW and JDA performed all wastewater monitoring activities including submission of samples for NGS, MRW supported data analysis and submission to the health department and manuscript revision, RLU served as a liaison between wastewater treatment plants and student research assistants, and EWA assisted in data analysis and manuscript revision.

## Acknowledgements

We thank MDHHS and MiNET for supporting wastewater collection, processing, and data analysis. We also thank the WWTP staff who provided samples every week during a pandemic. We thank CMU undergraduate assistants Justus Holben, Gabrielle Reau, Jessica Broach, Hamzah Khan, Jayde-Ann Taylor, Ashley Bergmooser, Kaitlyn Perry, Emily Rosema, Alexis Bruce, and Lauren Revord for their support. This is contribution number XXX of the Central Michigan University Institute for Great Lakes Research.

## Figure legends

**Supplementary Figure 1. Reconstructed ORFs in FASTA format**

**Supplementary Table 1. Total RNA-Seq reads present for each SARS-CoV-2 ORF**

