## Supplementary Figure 1 for "Wastewater sequencing from a rural community enables identification of widespread adaptive mutations in a SARS-CoV-2 Alpha variant"

Reconstructed ORFs in FASTA format

>NSP1

MESLVPGFNEKTHVQLILPVLQVRDVLVRGFGDSVEEVLSEARQHLKDGTCGLVEVEKGVLPQLEQPYVFIKRSDARTAPHGHVMVELVAELEGIQYGRSGETLGVLVPHVGEIPVAYRKVLLRKNGNKGAGGHSYGADLKSFDLGDELGTDPYEDFQENWNTKHSSGVTRELMRELNGG

>NSP2

AYTRYVDNNFCGPDGYPLECIKDLLARAGKASCTLSEQLDFIDTKRGVYCCREHEHEIAWYTERSEKSYELQTPFEIKLAKKFDTFNGECPNFVFPLNSIIKTIQPRVEKKKLDGFMGRIRSVYPVASPNECNQMCLSTLMKCDHCGETSWQTGDFVKATCEFCGTENLTKEGATTCGYLPQNAVVKIYCPACHNSEVGPEHSLAEYHNESGLKTILRKGGRTIAFGGCVFSYVGCHNKCAYWVPRASANIGCNHTGVVGEGSECLNDNLLEILQKEKVNINIVGDFKLNEEIAIILASFSASTSAFVETVKGLDYKAFKQIVESCGNFKVTKGKAKKGAWNIGEQKSILSPLYAFASEAARVVRSIFSRTLETAQNSVRVLQKAAITILDGISQYSLRLIDAMMFTSDLATNNLVVMAYITGGVVQLTSQWLTNIFGTVYEKLKPVLDWLEEKFKEGVEFLRDGWEIVKFISTCACEIVGGQIVTCAKEIKESVQTFFKLVNKFLALCADSIIIGGAKLKALNLGETFVTHSKGLYRKCVKSREETGLLMPLKAPKEIIFLEGETLPTEVLTEEVVLKTGDLQPLEQPTSEAVEAPLVGTPVCINGLMLLEIKDTEKYCALAPNMMVTNNTFTLKGG

>NSP3

APTKVTFGDDTVIEVQGYKSVNITFELDERIDKVLNEKCSAYTVELGTEVNEFACVVADAVIKTLQPVSELLTPLGIDLDEWSMATYYLFDESGEFKLASHMYCSFYPPDEDEEEGDCEEEEFEPLTQYEYGTEDDYQGKPLEFGATSAALQPEEEQEEDWLDDDSQQTVGQQDGSEDNQTTIIQTIVEVQPQLEMELTPVVQTIEVNSFSGYLKLTDNVYIKNADIVEEAKKVKPTVVVNAANVYLKHGGGVAGALNKATNNAMQVESDDYIATNGPLKVGGSCVLSGHNLAKHCLHVVGPNVNKGEDIQLLKSAYENFNQHEVLLAPLLSAGIFGADPIHSLRVCVDTVRTNVYLAVFDKNLYEKLVSSFLEMKSEKQVEQKIAEIPKEEVKPFITESKPSVEQRKQDDKKIKACVEEVTTTLEETKFLTENLLLYIDINGNLHPDSATLVSDIDITFLKKDAPYIVGDVVQEGVLTAVVIPTKKSGGTTEMLAKALRKVPTDNYITTYPGQGLNGYTVEEAKTVLKKCKSAFYILPSIISNEKQEILGTVSWNLREMLAHAEETRKLMPVCVETKAIVSTIQRKYKGIKIQEGVVDYGARFYFYTSKTTVASLINTLNDLNETLVTMPLGYVTHGLNLEEAARYMRSLKVPATVSVSSPDAVTAYNGYLTSSSKTPEEHFIETISLAGSYKDWSYSGQSTQLGIEFLKRGDKSVYYTSNPTTFYLDGEVITFDNLKTLLSLREVRTIKVFTTVDNINLHTQVVDMSMTYGQQFGPTYLDGADVTKIKPHNSHEGKTFYVLPNDDTLRVEAFEYYHTTDPSFLGRYMSALNHTKKWKYPQVNGLTSIKWADNNCYLATALLTLQQIELKFNPPALQDAYYRARAGEADNFCALILAYCNKTVGELGDVRETMSYLFQHANLDSCKRVLNVVCKTCGQQQTTLKGVEAVMYMGTLSYEQFKKGVQIPCTCGKQATQYLVQQESPFVMMSAPPAQYELKHGTFTCASEYTGNYQCGHYKHITSKETLYCIDGALLTKSSEYKGPITDVFYKENSYTTTIKPVTYKLDGVVCTEIDPKLDNYYKKDNSYFTEQPIDLVPNQPYPNASFDNFKFVCDNIKFADDLNQLTGYKKPASRELKVTFFPDLNGDVVAIDYKHYTPSFKKGAKLLHKPIVWHVNNATNKATYKPNTWCIRCLWSTKPVETSNSFDVLKSEDAQGMDNLACEDLKPVSEEVVENPTIQKDVLECNVKTTEVVGDIILKPANNSLKITEEVGHTDLMAAYVDNSSLTIKKPNELSIVLGLKTLATHGLAAVNSVPWDTIANYAKPFLNKVVSTTTNIVTRCLNRVCTNYMPYFFTLLLQLCTFTRSTNSRIKASMPTTIAKNTVKSVGKFCLEASFNYLKSPNFSKLINITIWFLLLSVCLGSLIYSTAALGVLMSNLGMPSYCTGYREGYLNSTNVTIATYCTGSIPCSVCLSGLDSLDTYPSLETIQITISSFKWDLTAFGLVAEWFLAYILFTRFFYVLGLAAIMQLFFSYFAVHFISNSWLMWLIINLVQMAPISAMVRMYIFFASFYYVWKSYVHVVDGCNSSTCMMCYKRNRATRVECTTIVNGVRRSFYVYANGGKGFCKLHNWNCVNCDTFCAGSTFISDEVARDLSLQFKRPINPTDQSSYIVDSVTVKNGSIHLYFDKAGQKTYERHSLSNFVNLDNLRANNTKGSLPINVIVFDGKSKCEESSAKSASVYYSQLMCQPILLLDQALVSDVGDSAEVAVKMFDAYVNTFSSTFNVPMEKLKTLVATAEAELAKNVSLDNVLSTFISAARQGFVDSDVETKDVVECLKLSHQFDIEVTGDSCNNYMLTYNKVENMTPRDLGACIDCSARHINAQVAKSHNIALIWNVKDFMSLSEQLRKQIRSAAKKNNLPFKLTCATTRQVVNVVTTKIALKGG

>NSP4

KIVNNWLKQLIKVTLVFLFVAAIFYLITPVHVISKHTDFSSEIIGYKAIDGGVTRDIASTDTCFANKHADFDTWFSQRGGSYTNDKACPLIAAVITREVGFVVPGLPGTILRTTNGDFLHFLPRVFSAVGNICYTPSKLIEYTDFATSACVLAAECTIFKDASGKPVPYCYDTNVLEGSFAYESLRPDTRYVLMDGSFIQFPNTYLEGSVRVVTTFGSEYCRHGTCERSEAGVCVSTSGRWVLNNDYYRSLPGVFCGVDAVNLLTNMFTPLIQPIGALDISASIVAGGIVAIVVTCLAYYFMRFRRAFGEYSHVVAFNTLLFLMSFTVLCLTPVYSFLPGVYSVIYLYLTFYLTNDVSFLAHIQWMVMFTPLVPFWITIAYIICISTKHFYWFFSNYLKRRVVFNGVSFSTFEEAALCTFLLNKEMYLKLRSDVLLPLTQYNRYLALYNKYKYFSGAMDTTSYREAACCHLAKALNDFSNSGSDVLYQPPQTSITSAVLQ

>NSP5-3CLpro

SGFRKMAFPSGKVEGCMVQVTCGTTTLNGLWLDDVVYCPRHVICTSEDMLNPNYEDLLIRKSNHNFLVQAGNVQLRVIGHSMQNCVLKLKVDTANPKTPKYKFVRIQPGQTFSVLACYNGSPSGVYQCAMRPNFTIKGSFLNGSCGSVGFNIDYDCVSFCYMHHMELPTGVHAGTDLEGNFYGPFVDRQTAQAAGTDTTITVNVLAWLYAAVINGDRWFLNRFTTTLNDFNLVAMRYNYEPLTQDHVDILGPLSAQTGIAVLDMCASLKELLQNGMNGRTILGSALLEDEFTPFDVVRQCSGVTFQ

>NSP6

SAVKRTIKGTHHWLLLTILTSLLVLVQSTQWSLFFFFYENAFLPFAMGIIAMSAFAMMFVKHKHAFLCLFLLPSLATVAYFNMVYMPASWVMRIMTWLDMVDTSLAFKLKDCVMYASAVVLLILMTARTVYDDGARRVWTLMNVLTLVYKVYYGNALDQAISMWALIISVTSNYSGVVTTVMVLARGIVFMCVEYCPIFFITGNTLQCIMLVYCFLGYFCTCYFGLFCLLNRYFRLTLGVYDYLVSTQEFRYMNSQGLLPPKNSIDAFKLNIKLLGVGGKPCIKVATVQ

>NSP7

SKMSDVKCTSVVLLSVLQQLRVESSSKLWAQCVQLHNDILLAKDTTEAFEKMVSLLSVLLSMQGAVDINKLCEEMLDNRATLQ

>NSP8

AIASEFSSLPSYAAFATAQEAYERAVANGDSEVVLKKLKKSLNVAKSEFDRDAAMQRKLEKMADQAMTQMYKQARSEDKRAKVTSAMQTMLFTMLRKLDNDALNNIINNARDGCVPLNIIPLTTAAKLMVVIPDYNTYKNTCDGTTFTYASALWEIQQVVDADSKIVQLSEISMDNSPNLAWPLIVTALRANSAVKLQ

>NSP9

NNELSPVALRQMSCAAGTTQTACTDDNALAYYNTTKGGRFVLALLSDLQDLKWARFPKSDGTGTIYTELEPPCRFVTDTPKGPKVKYLYFIKGLNNLNRGMVLGSLAATVRLQ

>NSP10

AGNATEVPANSTVLSFCAFAVDAAKAYKDYLASGGQPITNCVKMLCTHTGTGQAITVTPEANMDQESFGGASCCLYCRCHIDHPNPKGFCDLKGKYVQIPTTCANDPVGFTLKNTVCTVCGMWKGYGCSCDQLREPMLQ

>RdRp

REPMLQSADAQSFLNRVCGVSAARLTPCGTGTSTDVVYRAFDIYNDKVAGFAKFLKTNCCRFQEKDEDDNLIDSYFVVKRHTFSNYQHEETIYNLLKDCPAVAKHDFFKFRIDGDMVPHISRQRLTKYTMADLVYALRHFDEGNCDTLKEILVTYNCCDDDYFNKKDWYDFVENPDILRVYANLGERVRQALLKTVQFCDAMRNAGIVGVLTLDNQDLNGNWYDFGDFIQTTPGNGVPVVDSYYSLLMPILTLTRALTAESHVDTDLTKPYIKWDLLKYDFTEERLKLFDRYFKYWDQTYHPNCVNCLDDRCILHCANFNVLFSTVFPLTSFGPLVRKIFVFVVSTGYHFRELGVVHNQDVNLHSSRLSFKELLVYAADPAMHAASGNLLLDKRTTCFSVAALTNNVAFQTVKPGNFNKDFYDFAVSKGFFKEGSSVELKHFFFAQDGNAAISDYDYYRYNLPTMCDIRQLLFVVEVVDKYFDCYDGGCINANQVIVNNLDKSAGFPFNKWGKARLYYDSMSYEDQDALFAYTKRNVIPTITQMNLKYAISAKNRARTVAGVSICSTMTNRQFHQKLLKSIAATRGATVVIGTSKFYGGWHNMLKTVYSDVENPHLMGWDYPKCDRAMPNMLRIMASLVLARKHTTCCSLSHRFYRLANECAQVLSEMVMCGGSLYVKPGGTSSGDATTAYANSVFNICQAVTANVNALLSTDGNKIADKYVRNLQHRLYECLYRNRDVDTDFVNEFYAYLRKHFSMMILSDDAVVCFNSTYASQGLVASIKNFKSVLYYQNNVFMSEAKCWTETDLTKGPHEFCSQHTMLVKQGDDYVYLPYPDPSRILGAGCFVDDIVKTDGTLMIERFVSLAIDAYPLTKHPNQEYAAVFHLYLQYIRKLHDELTGHMLDMYSVMLTNDNNSRYWEPEFYEAMYTPHTVLQ

>NSP13

AVGACVLCNSQTSLRCGACIRRPFLCCKCCYDHVISTSHKLVLSVNPYVCNAPGCDVTDVTQLYLGGMSYYCKSHKPPISFPLCANGQVFGLYKNTCVGSDNVTDFNAIATCDWTNAGDYILANTCTERLKLFAAETLKATEETFKLSYGIATVREVLSDRELHLSWEVGKPRPPLNRNYVFTGYRVTKNSKVQIGEYTFEKGDYGDAVVYRGTTTYKLNVGDYFVLTSHTVMPLSAPTLVPQEHYVRITGLYPTLNISDEFSSNVANYQKVGMQKYSTLQGPPGTGKSHFAIGLALYYPSARIVYTACSHAAVDALCEKALKYLPIDKCSRIIPARARVECFDKFKTDKFKVNSTLEQYVFCTVNALPETTADIVVFDEISMATNYDLSVVNARLRAKHYVYIGDPAQLPAPRTLLTKGTLEPEYFNSVCRLMKTIGPDMFLGTCRRCPAEIVDTVSALVYDNRLKAHKDKSAQCFKMFYKGVITHDVSSAINRPQIGVVREFLTRNPAWRKAVFISPYNSQNAVASKILGLPTQTVDSSQGSEYDYVIFTQTTETAHSCNVNRFNVAITRAKVGILCIMSDRDLYDKLQFTSLEIPRNVATLQ

>NSP14

AENVTGLFKDCSKVITGLHPTQAPTHLSVDTKFKTEGLCVDIPGIPKDMTYRRLISMMGFKMNYQVNGYPNMFITREEAIRHVRAWIGFDVEGCHATREAVGTNLPLQLGFSTGVNLVAVPTGYVDTPNNTDFSRVSAKPPPGDQFKHLIPLMYKGLPWNVVRIKIVQMLSDTLKNLSDRVVFVLWAHGFELTSMKYFVKIGPERTCCLCDRRATCFSTASDTYACWHHSIGFDYVYNPFMIDVQQWGFTGNLQSNHDLYCQVHGNAHVASCDAIMTRCLAVHECFVKRVDWTIEYPIIGDELKINAACRKVQHMVVKAALLADKFPVLHDIGNPKAIKCVPQADVEWKFYDAQPCSDKAYKIEELFYSYATHSDKFTDGVCLFWNCNVDRYPANSIVCRFDTRVLSNLNLPGCDGGSLYVNKHAFHTPAFDKSAFVNLKQLPFFYYSDSPCESHGKQVVSDIDYVPLKSATCITRCNLGGAVCRHHANEYRLYLDAYNMMISAGFSLWVYKQFDTYNLWNTFTRLQ

>NSP15

SLENVAFNVVNKGHFDGQQGEVPVSIINNTVYTKVDGVDVELFENKTTLPVNVAFELWAKRNIKPVPEVKILNNLGVDIAANTVIWDYKRDAPAHISTIGVCSMTDIAKKPTETICAPLTVFFDGRVDGQVDLFRNARNGVLITEGSVKGLQPSVGPKQASLNGVTLIGEAVKTQFNYYKKVDGVVQQLPETYFTQSRNLQEFKPRSQMEIDFLELAMDEFIERYKLEGYAFEHIVYGDFSHSQLGGLHLLIGLAKRFKKSPLELEDFIPMDSTVKNYFITDAQTGSSKCVCSVIDLLLDDFVEIIKSQDLSVVSKVVKVTIDYTEISFMLWCKDGHVETFYPKLQ

>NSP16

SSQAWQPGVAMPNLYKMQRMLLEKCDLQNYGDSATLPKGIMMNVAKYTQLCQYLNTLTLAVPYNMRVIHFGAGSDKGVAPGTAVLRQWLPTGTLLVDSDLNDFVSDADSTLIGDCATVHTANKWDLIISDMYDPKTKNVTKENDSKEGFFTYICGFIQQKLALGGSVAIKITEHSWNADLYKLMGHFAWWTAFVTNVNASSSEAFLIGCNYLGKPREQIDGYVMHANYIFWRNTNPIQLSSYSLFDMSKFPLKLRGTAVMSLKEGQINDMILSLLSKGRLIIRENNRVVISSDVLVNN

>ORF3a

MDLFMRIFTIGTVTLKQGEIKDATPSDFVRATATKPIQASLPFGWLIVGVALLAVFQSASKIITLKKRWQLALSKGVHFVCNLLLLFVTVYSHLLLVAAGLDAPFLYLYALVYFLQSINFVRIIMRLWLCWKCRSKNPLLYDANYFLCWHTNCYDYCIPYNSVTSSIVITSRDGTTSPISEHDYQIGGYTEKWESGVKDCVVLHSYFTSDYYQLYSTQLSTDTGVEHVTFFIYNKIVDEPEEHVQIHTIDGSSGVVNPVKEPIYDEPTTTTSVPL

>Envelope

MYSFVSEETGTLIVNSVLLFLAFVVFLLVTLAILTALRLCAYCCNIVNVSLVKPSFYVYSRVKNLNSSRVPDLLV

>Membrane

MANSNGTITVEELKKLLEHWNLVIGFLFLTWICLLQFAYANRNRFLYIIKLIFLWLLWPVTLACFVLAAVYRINWITGGIAIAMACLVGLMWLSYFIASFRLFARTRSMWSFNPETNILLNVPLHGTILTRPLLESELVIGAVILRGHLRIAGHHLGRCDIKDLPKEITVATSRTLSYYKLGASQRVAGDSGFAAYSRYRIGNYKLNTDHSSSSDNIALLVQ

>ORF6

MFHLVDFQVTIAEILLIIMRTFKVSIWNLDYIINLIIKNLSKSLTENKYSQLDEEQPMEIE

>ORF7a

MKIILFLALITLATCELYHYQECVRGTTVLLKEPCSSGTYEGNSPFHPLADNKFALTCFSTQFAFACPDGVKHVYQLRARSVSPKLFIRQEEVQELYSPIFLIVAAIVFITLCFTLKRKIE

>ORF7b

MIELSLIDFYLCFLAFLLFLVLIMLIIFWFSLELQDHNETCHV

>ORF8

MKFLVFLGIITTVAAFHQECSLQSCTHQPYVVDDPCPIHFYSKWYIRVGAIKSAPLIELCVDEAGSKSPIQCIDIGNYTVSCLPFTINCQEPKLGSLVVRCSFYEDFLEYHDVRVVLDFI

>Nucleocapsid

MSDNGPQDQRNAPRITFGGPSDSTGSNQNGERSGARPKQRRPQGLPNNTASWFTALTQHGKEDLKFPRGQGVPINTNSSPDDQIGYYRRATRRIRGGDGKMKDLSPRWYFYYLGTGPEAGLPYGANKDGIIWVATEGALNTPKDHIGTRNPANNAAIVLQLPQGTTLPKGFYAEGSRGGSQASSRSSSRSRNSSRNSTPGSSRGTSPARMAGNGGDAALALLLLDRLNQLESKMSGKGQQQQGQTVTKKSAAEASKKPRQKRTATKAYNVTQAFGRRGPEQTQGNFGDQELIRQGTDYKHWPQIAQFAPSASAFFGMSRIGMEVTPSGTWLTYTGAIKLDDKDPNFKDQVILLNKHIDAYKTFPPTEPKKDKKKKADETQALPQRQKKQQTVTLLPAADLDDFSKQLQQSMSSADSTQA

>ORF10

MGYINVFAFPFTIYSLLLCRMNSRNYIAQVDVVNFNLT
