## Supplementary Table 1 for "Wastewater sequencing from a rural community enables identification of widespread adaptive mutations in a SARS-CoV-2 Alpha variant"

Supplementary Table 1. Total RNA-Seq reads present for each SARS-CoV-2 ORF

| ORF | 10/26/2021 | 11/9/2021 | 9/12/2022 | 3/13/2023 | 3/27/2023 | 4/24/2023 | 5/1/2024 |
| --- | --- | --- | --- | --- | --- | --- | --- |
| Nsp1 | 209800 | 54980 | 41452 | 42616 | 75064 | 55170 | 47560 |
| Nsp2 | 20258 | 86300 | 64384 | 108024 | 39338 | 64276 | 149306 |
| Nsp3 | 82106 | 156766 | 140460 | 215204 | 93880 | 114824 | 398662 |
| Nsp4 | 25374 | 30990 | 33054 | 65438 | 11026 | 27832 | 66442 |
| Nsp5 | 18568 | 51018 | 59714 | 48180 | 35350 | 34714 | 90384 |
| Nsp6 | 5128 | 46436 | 28456 | 64276 | 16282 | 33470 | 72942 |
| Nsp7 | 8302 | 14044 | 4428 | 13110 | 3950 | 10046 | 12156 |
| Nsp8 | 23860 | 37972 | 18900 | 46478 | 11962 | 17414 | 48258 |
| Nsp9 | 1698 | 17566 | 18596 | 39140 | 27654 | 9000 | 40866 |
| Nsp10 | 34494 | 47696 | 52908 | 99674 | 46310 | 15842 | 90196 |
| RdRp | 153286 | 157946 | 75812 | 174802 | 32754 | 51030 | 222774 |
| Nsp15 | 1780 | 20414 | 13782 | 20172 | 4632 | 22378 | 48422 |
| Nsp16 | 746 | 13202 | 11490 | 15216 | 4938 | 10402 | 25376 |
| ORF3a | 2104 | 48900 | 28548 | 49744 | 22070 | 34572 | 102914 |
| M | 2222 | 36410 | 18364 | 25812 | 20910 | 34240 | 53452 |
| ORF6 | 64 | 16570 | 11984 | 7610 | 14448 | 5948 | 21916 |
| ORF7a | 754 | 9826 | 4216 | 7074 | 5620 | 10716 | 18644 |
| ORF7b | 54 | 3166 | 5444 | 11186 | 9422 | 4346 | 6568 |
| N | 228036 | 137820 | 88620 | 332662 | 107064 | 66860 | 161528 |
